# Investigating Automatic Speech Emotion Recognition for Children with Autism Spectrum Disorder in interactive intervention sessions with the social robot Kaspar

**DOI:** 10.1101/2022.02.24.22271443

**Authors:** Manuel Milling, Katrin D. Bartl-Pokorny, Björn W. Schuller

## Abstract

In this contribution, we present the analyses of vocalisation data recorded in the first observation round of the European Commission’s Erasmus Plus project “EMBOA, Affective loop in Socially Assistive Robotics as an intervention tool for children with autism”. In total, the project partners recorded data in 112 robot-supported intervention sessions for children with autism spectrum disorder. Audio data were recorded using the internal and lapel microphone of the H4n Pro Recorder. To analyse the data, we first utilise a child voice activity detection (VAD) system in order to extract child vocalisations from the raw audio data. For each child, session, and microphone, we provide the total time child vocalisations were detected. Next, we compare the results of two different implementations for valence- and arousal-based speech emotion recognition, thereby processing (1) the child vocalisations detected by the VAD and (2) the total recorded audio material. We provide average valence and arousal values for each session and condition. Finally, we discuss challenges and limitations of child voice detection and audio-based emotion recognition in robot-supported intervention settings.

## 1. Introduction

Children with autism spectrum disorder (ASD; OMIM 209850) have problems in social interactions and show restricted, repetitive patterns of behaviour, interests, or activities (American Psychiatric Association, 2013). Previous studies revealed that children with ASD usually prefer interactions with human-like looking robots to interactions with humans and therefore benefit from robot-based interventions in order to improve their socio-communicative skills (Bartl-Pokorny et al., 2021). To date, robots used in intervention settings with children with ASD need lots of manual control by therapists as they are not able to automatically and in real time respond to a child’s needs and emotional states. To overcome these limitations of social robots in the future, current research approaches focus on the implementation of automatic emotion recognition systems.

The EU Erasmus+ project “Affective loop in Socially Assistive Robotics as an intervention tool for children with autism” (https://emboa.eu), conducted by an international and multidisciplinary consortium of researchers from Gdansk University of Technology (GUT; Poland), University of Hertfordshire (UH; United Kingdom), Istanbul Teknik Universitesi (ITU; Turkey), Yeditepe University Vakif (YT; Turkey), the Macedonian Association for Applied Psychology (MAAP; North Macedonia), and University of Augsburg (UA; Germany), focuses on the practical evaluation of the use of state-of-the-art emotion recognition technologies in robot-supported interventions and aims to develop guidelines for the application of such technologies in intervention settings.

The project team collects data from multiple observation channels (e.g., vocalisations, eye movements, facial expressions, physiological parameters) during intervention sessions in which children with ASD are interacting with the social robot Kaspar (https://www.herts.ac.uk/kaspar). Based on these data, the researchers aim to evaluate each observation channel in terms of its information gain for automatic emotion recognition. This report focuses on the analyses of vocalisation data recorded in the first observation round.

The report is organised as follows: section 2 describes the available audio data, section 3 provides a description of the utilised child voice activity detection (VAD) system and results on detected vocalisations, section 4 describes the applied speech emotion recognition model (SER) and the detected valence and arousal values. Section 5 sums up the results and discusses revealed challenges and limitations of automatic child voice detection and audio-based emotion recognition in robot-supported intervention settings.

## 2. Methods

### 2.1. Audio data

The EMBOA team received ethics approval by Gdansk University of Technology/Poland for conducting the study. The inclusion criteria for the present study were: (1) Children were diagnosed with ASD by a mental health professional in the country of living, (2) children were recruited in institutions in the partner countries North Macedonia, Turkey, Poland, and UK, (3) children had already received treatments for ASD in their caregiving institutions or were newly diagnosed and had not received treatments before inclusion in the study, (4) children were between 2 and 12 years old, (4) the children’s legal guardians signed an informed consent form for participation in the study. As there were no further inclusion criteria defined, the sample was heterogeneous regarding amongst others gender, exact age, language skills, and comorbidities. There were neither clinical assessments nor follow-up investigations done with the children in the framework of the present study.

The participating children with ASD were recorded during intervention sessions that were supported by the social robot Kaspar. During these interventions, at least one researcher and one parent were together with the child and the robot in a room. The audio data were recorded from the partners at GUT/Poland, ITU-YU/Turkey, MAAP/North Macedonia, and UH/United Kingdom using the internal and lapel microphone of the H4n Pro Recorder. For most of the recorded robot-supported intervention sessions, both the recordings from the internal and from the lapel microphone are available. Recordings with the internal microphone of four sessions of MAAP/North Macedonia and recordings with the lapel microphone of all fourteen sessions of ITU-YU/Turkey were excluded from analysis due to insufficient audio quality. Table 1 provides an overview of the final set of analysable audio recordings. The children with ASD were recorded in up to eleven intervention sessions, as listed in Table 2. The minimum number of recorded intervention sessions should be 2 for all children; the actual number of recorded sessions depended on the individual therapeutic needs of the children, the family’s willingness to continue participation, and the achievement of the project goals regarding sample size. The children recorded by GUT/Poland were 6 years old, the children recorded by ITU-YU/Turkey were between 6 and 10 years old, the children recorded by MAAP/North Macedonia were between 2 and 6 years old, and the children recorded by UH/United Kingdom were 11 to 12 years old. All data were recorded in the years 2020 and 2021. The number of included children was in conformity with the EMBOA project protocol approved by the funding agency.

**Table 1:**
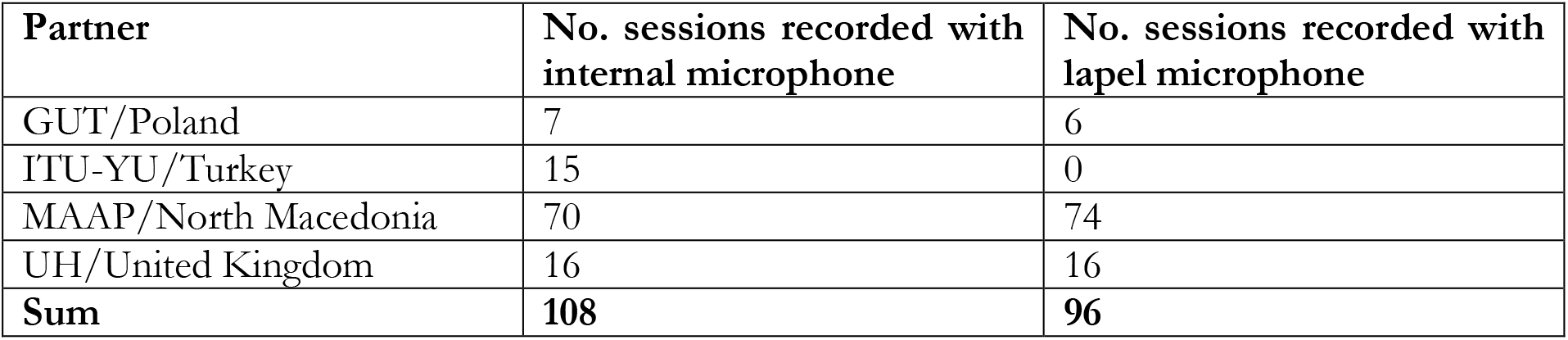
Audio data included in analysis. GUT = Gdansk University of Technology, Poland; ITU-YU = Istanbul Teknik Universitesi and Yeditepe University Vakif, Turkey; MAAP = Macedonian Association for Applied Psychology, North Macedonia; UH = University of Hertfordshire, United Kingdom.

| **Partner** | **No. sessions recorded with internal microphone** | **No. sessions recorded with lapel microphone** |
| --- | --- | --- |
| GUT/Poland | 7 | 6 |
| ITU-YU/Turkey | 15 | 0 |
| MAAP/North Macedonia | 70 | 74 |
| UH/United Kingdom | 16 | 16 |
| **Sum** | **108** | **96** |

**Table 2:**
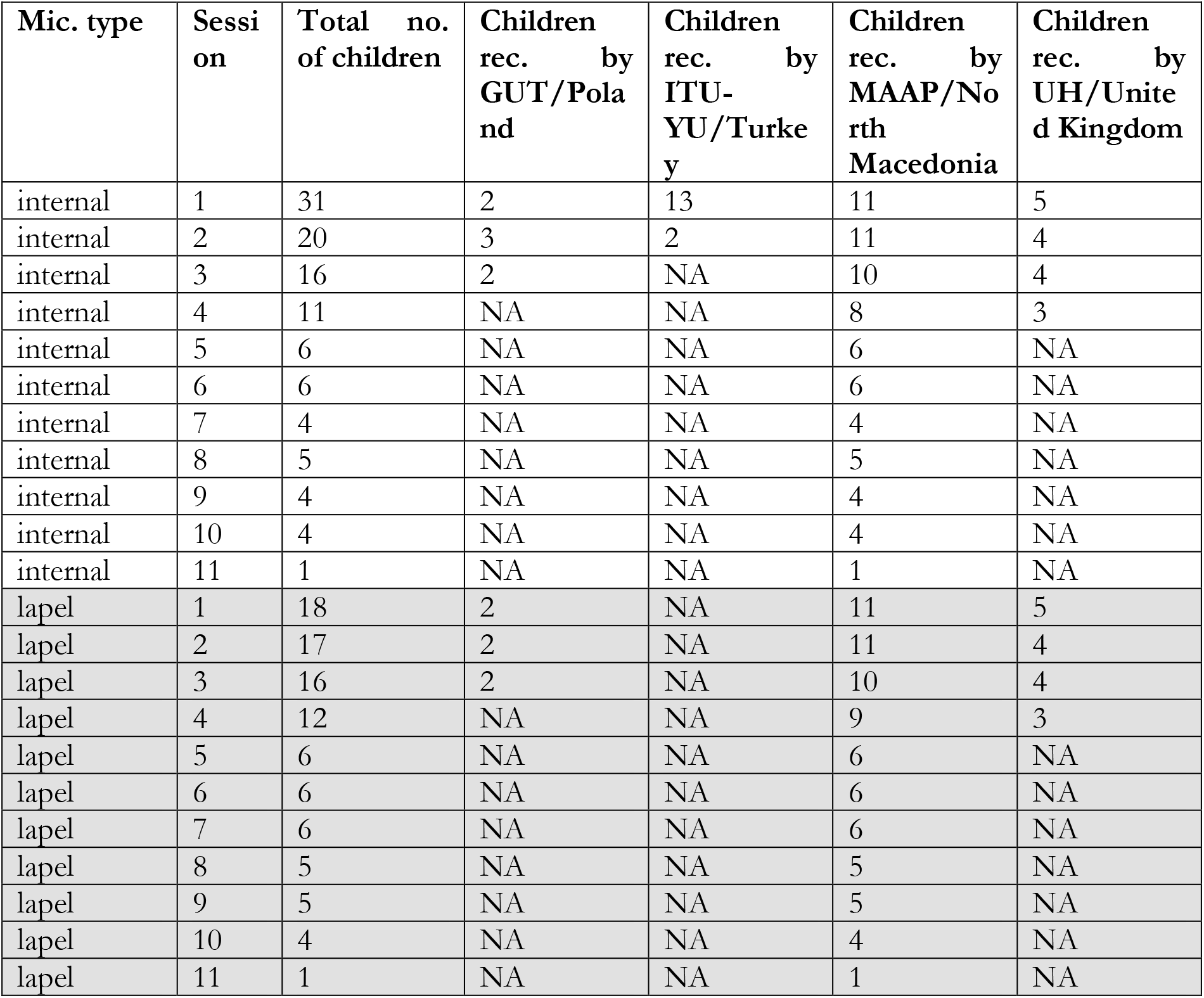
Availability of analysable audio data for each microphone type and session. GUT = Gdansk University of Technology, Poland; ITU-YU = Istanbul Teknik Universitesi and Yeditepe University Vakif, Turkey; MAAP = Macedonian Association for Applied Psychology, North Macedonia; Mic. = microphone; NA = not available/insufficient audio quality, no. = number, rec. = recorded, UH = University of Hertfordshire, United Kingdom.

We are aware that certain systematic differences between the data recorded from the different partners such as age, mother tongue, individual language skills of the children, room acoustics, etc. may cause bias. Therefore, we report quantitative results separately for individual children and/or participating sites and discuss qualitative differences in the data.

### 2.2. Child voice activity detection

Following Milling et al. (2022), we apply a deep learning-based voice activity detection (VAD) system specifically trained for vocalisations of children with ASD to extract child vocalisations, as we expect the child vocalisations to contain the most information about the children’s affective states. The used VAD system has been trained and evaluated on data recorded under similar conditions as the EMBOA data, namely in the framework of the European Commission’s DE-ENIGMA project (Shen et al., 2018; https://cordis.europa.eu/project/id/688835; https://de-enigma.eu). For the intervention sessions in DE-ENIGMA, the robot Zeno was used. The model was trained on audio data from the British study arm of the project containing audio data from 84 robot-supported intervention sessions of 25 English-speaking children with ASD from the United Kingdom, having a total duration of more than 17 hours. For the experiments reported by Milling et al. (2022), the data was partitioned into train, development, and test partitions in a speaker-independent way. In total, this data contains more than 2 hours of annotated child vocalisations (partially overlapping with other vocalisations). The system was trained to predict the probability of a child vocalisation being present in a given 10-millisecond-long audio frame based on so-called low-level descriptors (LLDs) of the said audio frame. LLDs are a common set of characteristics, which can be extracted from an audio signal, including features such as fundamental frequency (F0), variations in the volume (shimmer), and variations in the fundamental frequency (jitter). The deep learning approach consists of a two-layer bi-directional long short-term memory (Bi-LSTM) recurrent neural network (RNN), followed by a dense layer, which is designed to allow information flow for several consecutive audio frames. Different thresholds for the predicted probability of the system allow for the adjustment of trade-off between sensitivity and specificity. For inference, a threshold corresponding to the equal-error-rate (EER) was chosen, i.e., equal values of true positive rate and false positive rate. Finally, the VAD system summarises frame-level predictions with a detection event if the EER threshold is reached for at least 0.25 s of a 1-s-chunk. More details on the VAD are given in Milling et al. (2022). The child VAD system has a reported EER of 0.381 and an area-under-the-curve of 0.662 on the DE-ENIGMA corpus. However, even though the recording conditions of both projects appear rather similar to each other, it is not clear whether this performance translates to the EMBOA data, as a lack of systematic vocalisation annotations prevents a thorough evaluation of the model.

### 2.3. Speech emotion recognition

In order to estimate emotions from speech we use a deep learning model developed, trained, and evaluated by Milling et al. (2022), which is again based on the same DE-ENIGMA data and partitioning of British children, as described for the VAD component. The models are designed for a continuous speech emotion recognition (SER) task and we apply correspondingly trained models for two types of tasks: 1) each 1 s chunk from the original audio recording is used for the continuous SER task and 2) only 1 s chunks, for which a child vocalisation was detected, are being used for the continuous SER task. Within each task, for each relevant audio chunk, we then extract 88 functional features of the extended Geneva Minimalistic Acoustic Parameter Set (eGeMAPS; Eyben et al., 2015) from 1 s audio chunks. The features are then fed into a model consisting of two RNN layers with LSTM cells and a hidden layer size of 128 units, a feed-forward layer with 128 neurons, a rectified linear unit (ReLU) activation and a dropout rate of 0.3, and a final feed-forward layer with a single neuron, predicting the valence values (and arousal values, respectively). Reported performances of the models achieve up to 0.168 concordance correlation coefficient (CCC) on the test partition of the DE-ENIGMA corpus. More details on the SER system and performance values can be found in Milling et al. (2022).

## 3. Results

### 3.1. Detected child vocalisations

A general overview of detected child vocalisations is provided in Table 3. Table 4 groups child voice activity detections by the session number in a consecutive series of sessions with partly identical children. Detailed session-wise results can be found in Table A.1. of the annexe. The proportion of time with detected child vocalisations per session is similar in the internal and lapel microphone condition (Table 3). The proportion of time with detected child vocalisations is similar for each of the 10 analysed sessions for children recorded by MAAP/North Macedonia (Table 4); we can neither observe an obvious increase nor decrease in terms of detected vocalisations with the children’s increasing experience with robot-supported intervention.

**Table 3:**
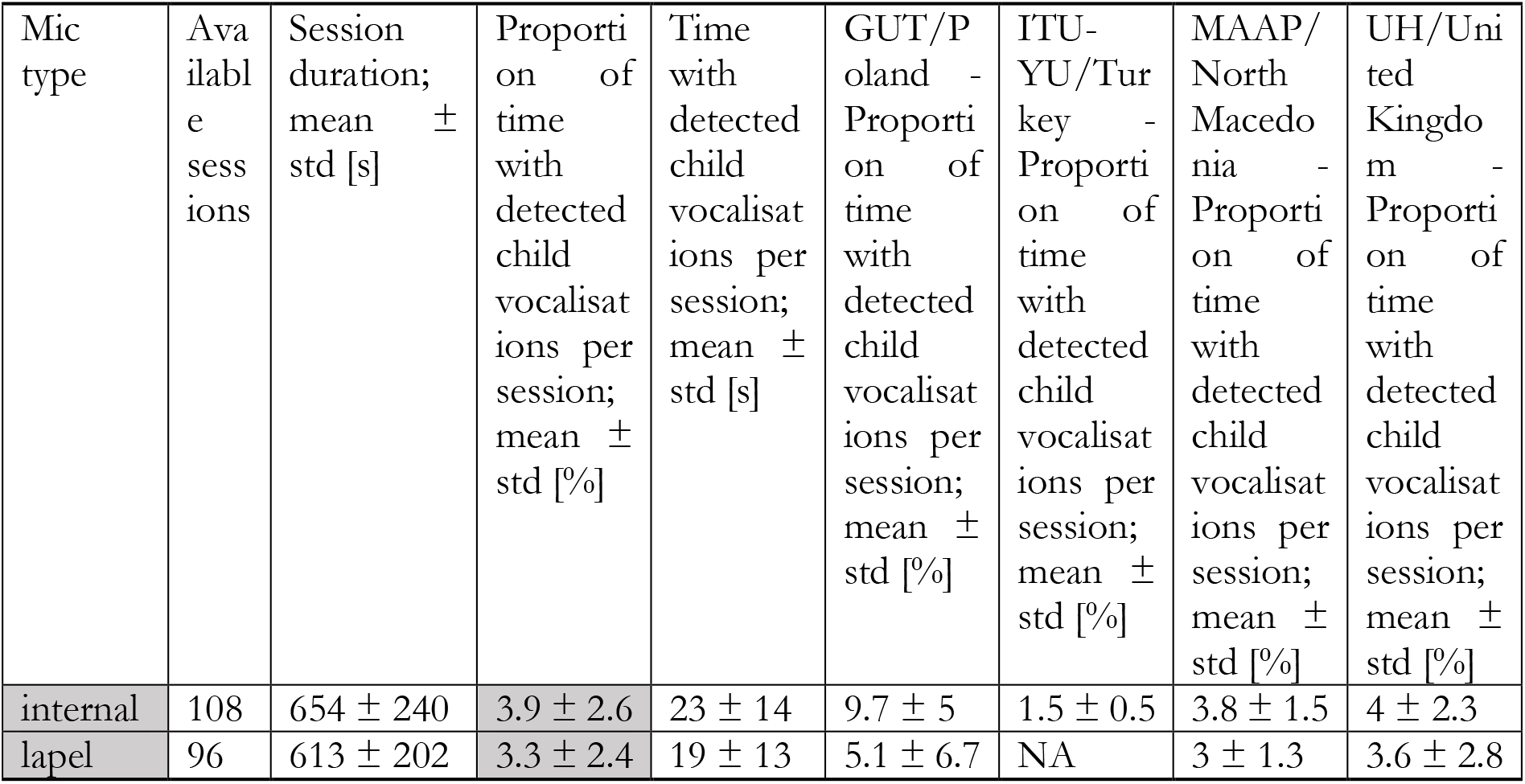
Overview of results on child voice activity detection. GUT = Gdansk University of Technology, Poland; ITU-YU = Istanbul Teknik Universitesi and Yeditepe University Vakif, Turkey; MAAP = Macedonian Association for Applied Psychology, North Macedonia; UH = University of Hertfordshire, United Kingdom.

| Mic type | Available sessions | Session duration; mean ± std [s] | Proportion of time with detected child vocalisations per session; mean ± std [%] | Time with detected child vocalisations per session; mean ± std [s] | GUT/Poland - Proportion of time with detected child vocalisations per session; mean ± std [%] | ITU-YU/Turkey - Proportion of time with detected child vocalisations per session; mean ± std [%] | MAAP/North Macedonia - Proportion of time with detected child vocalisations per session; mean ± std [%] | UH/United Kingdom - Proportion of time with detected child vocalisations per session; mean ± std [%] |
| --- | --- | --- | --- | --- | --- | --- | --- | --- |
| internal | 108 | 654 ± 240 | 3.9 ± 2.6 | 23 ± 14 | 9.7 ± 5 | 1.5 ± 0.5 | 3.8 ± 1.5 | 4 ± 2.3 |
| lapel | 96 | 613 ± 202 | 3.3 ± 2.4 | 19 ± 13 | 5.1 ± 6.7 | NA | 3 ± 1.3 | 3.6 ± 2.8 |

**Table 4:** Results on child voice activity detection for MAAP/North Macedonia data recorded with the internal microphone with respect to the session number in a consecutive series of sessions.

| Session No. | Available children | Session duration; mean ± std [s] | Proportion of time with detected child vocalisations; mean ± std [%] | Time with detected child vocalisations; mean ± std [s] |
| --- | --- | --- | --- | --- |
| 1 | 11 | 568 ± 254 | 3.6 ± 1.8 | 19 ± 10 |
| 2 | 11 | 577 ± 152 | 4.2 ± 1.2 | 24 ± 9 |
| 3 | 10 | 585 ± 80 | 3.5 ± 1.0 | 21 ± 7 |
| 4 | 8 | 776 ± 335 | 2.5 ± 0.8 | 18 ± 4 |
| 5 | 6 | 621 ± 113 | 4.6 ± 1.6 | 29 ± 13 |
| 6 | 6 | 663 ± 85 | 4.5 ± 1.2 | 30 ± 11 |
| 7 | 4 | 694 ± 46 | 3 ± 0.5 | 21 ± 4 |
| 8 | 5 | 619 ± 100 | 4 ± 2.2 | 25 ± 15 |
| 9 | 4 | 644 ± 108 | 4.7 ± 1.1 | 31 ± 11 |
| 10 | 4 | 674 ± 77 | 4.2 ± 1.3 | 28 ± 9 |

### 3.2. Predicted valence and arousal values

We apply the SER model with and without previous VAD-based pre-processing, as introduced in section 2.3, on each recorded session. Session-wise averaged emotion predictions are reported in Table A.2 of the annexe and are further discussed below.

## 4. Discussion

The aim of the present study was to investigate the applicability of existing SER approaches in robot-supported intervention sessions with children with ASD. For this, we deployed a publicly available pipeline of a child VAD and a SER model, both specifically trained for children with ASD. Both the VAD system and the SER system were developed based on data from the DE-ENIGMA project that also focused on robot-supported intervention for children with ASD.

For the present study, the VAD system raises a – partially limited – amount of detection events for the data at hand. The most impactful limitation of our study is that we cannot evaluate our models on the investigated data, collected within the EMBOA project, as annotations for vocalisations and emotions are lacking. This hampers the investigation of potential effects of different languages, different ages, background noise, etc. on voice detection. However, some observations can be noted based on the raw predictions of the network. First, the amount of detected events depends on the utilised microphone. In general, it seems that the internal microphone leads to a larger amount of detection events compared to the lapel microphone (i.e., 3.9 ± 2.6 vs 3.3 ± 2.4; Table 3), which was surprising as we initially hypothesised that the lapel microphone recordings would allow us to detect more vocalisations compared to the internal microphone recordings. For GUT/Poland data, actually the opposite was the case: The VAD system detected longer child vocalisation intervals in the internal microphone recordings compared to the lapel microphone recordings.

In an unstructured, exemplary analysis of the recording based on manual listening, we noticed that in the internal microphone recordings the voices of the other people in the room were considerably softer than the child’s voice. In comparison, the voices of the robot and the researcher as well as background noise in general was much louder in the lapel microphone recordings which is a potential reason for fewer detected child vocalisation intervals. When listening to parts of the audio recordings and comparing them with the automatically detected child vocalisation times, we noticed a number of further challenges for the VAD system: In some of the recordings, the audio recording levels of the microphone were too low which seems to correlate with no or only very few detected child vocalisations. As automatic emotion recognition cannot benefit from such recordings, future studies need to carefully check the microphone settings before recording robot-supported intervention sessions. In some cases, the microphone position resulted in further difficulties; e.g., some of the children did not accept the lapel microphone on their clothes and sometimes the clothes or table where the microphones were positioned on produced noise, which was partly louder than the child’s voice and seems to lower the detected child vocalisation time. In some of the recordings, potentially confounding factors related to room acoustics (e.g., ITU-YU/Turkey data were partly recorded in a room with long reverberation time) and background noise (e.g., in some of the MAAP/North Macedonia recordings, sound originating from chairs, tables, and doors is hearable) occurred. Overall, detected child vocalisation times seemed to be shorter in recordings with more people in a room compared to recordings with fewer overlay of voices (e.g., MAAP/North Macedonia data compared to GUT/Poland data). We also noticed that an apparent challenge for the VAD system is the detection of child vocalisation in the form of crying, which might be related to the limited number of crying instances in the original data. Similar observations can be reported for imitation of animal sounds and singing songs. Surprisingly, the VAD system detected relatively few child vocalisation events in the UH/United Kingdom recordings despite the children’s seemingly advanced language skills. It needs to be mentioned here that the children recorded by UH/United Kingdom were older than most of the children included in the DE-ENIGMA training material, which might have a negative effect on the child-specific VAD system.

As suggested by Milling et al. (2022), the performance of VAD systems can have an impact on the estimation of affective states based on audio recordings. This hypothesis is supported by the clear differences in valence and arousal predictions in Table 5, when comparing the average absolute difference between VAD-based SER predictions and the non-VAD-based SER predictions, in particular for the arousal case. It can further be reported that similarly to the VAD task, the choice of microphones seems to have an additional impact on the SER predictions, even though the observed effect is smaller compared to the differences caused by the VAD-based pre-processing.

**Table 5:** Average absolute deviation (*µ*) and standard deviation of absolute deviations (std) of affect prediction derived from Table A.1. Results are based on the same sessions respectively and compare pre-processing with (VAD-based) and without (non-VAD-based) a VAD system (of the identical microphone recording), as well as recordings from the lapel and the internal microphone (with identical VAD-based or non-VAD-based pre-processing).

| Comparison | Arousal ($\mu \pm \text{std}$) | Valence ($\mu \pm \text{std}$) |
| --- | --- | --- |
| VAD-based vs. non-VAD-based | $0.208 \pm 0.113$ | $0.055 \pm 0.045$ |
| internal vs. lapel | $0.086 \pm 0.068$ | $0.047 \pm 0.056$ |

However, the expressiveness of the SER models for the investigated data set has to be interpreted with caution as only some children in the EMBOA project have the same language and cultural background as the children based on which the model was trained, which potentially hinders the generalisation capabilities of said models. Additionally, the children in the EMBOA project were on average older than the children who provided the training data. To improve the reliability of the VAD and SER systems, the existing models, which were trained on the DE-ENIGMA data set, may be adapted to the EMBOA recording settings (e.g., acoustic room conditions, microphone type, specific age and language of children, etc.) based on annotated parts of the EMBOA data. Note that all reported observations on data quality and corresponding impact on model performance in this section rely on preliminary analyses and have not been quantitatively evaluated due to a lack of systematic annotations. Still, we gained a number of important insights with regard to VAD and SER that will help to formulate guidelines for the use of audio-based emotion recognition techniques in robot-supported intervention settings. Such guidelines will allow future studies to optimise their methodologies with regard to an improved audio-based automatic recognition of children’s emotional states in intervention settings. This will in turn help the development towards a social robot, which is capable of automatically reacting to a child’s current needs and could therefore enhance the child’s motivation to interact with the robot, potentially resulting in positive influences on the intervention success for children with ASD. A prerequisite for this will further be more robust emotion recognition models, being available for different language, culture, gender, and age groups.

## Supporting information

STROBE checklist

## Data Availability

Data used for the present study are speech data, i.e. personal data. The participants did not agree to share these data with people outside the EMBOA project team.

## Acknowledgements

We are very grateful to all children, families, and healthcare professionals for their participation in our study. We want to thank all project partners for data acquisition. Furthermore, we thank Alice Baird and Shuo Liu for consultancy in data analysis.

This publication was supported in part by the European Commission’s Erasmus Plus project “EMBOA, Affective loop in Socially Assistive Robotics as an intervention tool for children with autism”, contract no 2019-1-PL01-KA203-065096.

The European Commission’s support for the production of this publication does not constitute an endorsement of the contents which reflect the views only of the authors, and the Commission cannot be held responsible for any use which may be made of the information contained therein.

This report is distributed free of charge under Creative Commons License CC BY.

## Annexe

**Table A.1:** Detailed session-wise results on child voice activity detection. GUT = Gdansk University of Technology, Poland; ITU-YU = Istanbul Teknik Universitesi and Yeditepe University Vakif, Turkey; MAAP = Macedonian Association for Applied Psychology, North Macedonia; UH = University of Hertfordshire, United Kingdom.

| ID | Total recording length [s] | Detection proportion | Total detection time [s] |
| --- | --- | --- | --- |
| GUT-C02-S01-internal | 5 | 0.200 | 1 |
| GUT-C02-S01-lapel | 5 | 0.200 | 1 |
| GUT-C03-S01-internal | 35 | 0.114 | 4 |
| GUT-C03-S01-lapel | 35 | 0.029 | 1 |
| GUT-C01-S02-internal | 802 | 0.042 | 34 |
| GUT-C02-S02-internal | 580 | 0.070 | 41 |
| GUT-C02-S02-lapel | 580 | 0.017 | 10 |
| GUT-C03-S02-internal | 824 | 0.078 | 64 |
| GUT-C03-S02-lapel | 824 | 0.017 | 14 |
| GUT-C01-S03-internal | 805 | 0.053 | 43 |
| GUT-C01-S03-lapel | 805 | 0.016 | 13 |
| GUT-C02-S03-internal | 608 | 0.120 | 73 |
| GUT-C02-S03-lapel | 608 | 0.025 | 15 |
| ITU-YU-C01-S01-internal | 1814 | 0.017 | 32 |
| ITU-YU-C02-S01-internal | 821 | 0.018 | 15 |
| ITU-YU-C03-S01-internal | 605 | 0.026 | 16 |
| ITU-YU-C04-S01-internal | 644 | 0.020 | 13 |
| ITU-YU-C01-S02-internal | 937 | 0.018 | 17 |
| ITU-YU-C03-S02-internal | 752 | 0.015 | 11 |
| ITU-YU-C05-S01-internal | 713 | 0.022 | 16 |
| ITU-YU-C06-S01-internal | 933 | 0.014 | 13 |
| ITU-YU-C07-S01-internal | 997 | 0.006 | 6 |
| ITU-YU-C08-S01-internal | 713 | 0.011 | 8 |
| ITU-YU-C09-S01-internal | 832 | 0.010 | 8 |
| ITU-YU-C10-S01-internal | 1058 | 0.012 | 13 |
| ITU-YU-C11-S01-internal | 897 | 0.007 | 6 |
| ITU-YU-C12-S01-internal | 902 | 0.016 | 14 |
| ITU-YU-C13-S01-internal | 1033 | 0.011 | 11 |
| MAAP-C01-S01-internal | 885 | 0.023 | 20 |
| MAAP-C01-S01-lapel | 885 | 0.029 | 26 |
| MAAP-C01-S02-internal | 656 | 0.050 | 33 |
| MAAP-C01-S02-lapel | 656 | 0.038 | 25 |
| MAAP-C01-S03-internal | 665 | 0.054 | 36 |
| MAAP-C01-S03-lapel | 665 | 0.059 | 39 |
| MAAP-C01-S04-internal | 515 | 0.025 | 13 |
| MAAP-C01-S04-lapel | 515 | 0.027 | 14 |
| MAAP-C02-S01-internal | 277 | 0.014 | 4 |
| MAAP-C02-S01-lapel | 277 | 0.040 | 11 |
| MAAP-C02-S02-internal | 436 | 0.023 | 10 |
| MAAP-C02-S02-lapel | 436 | 0.014 | 6 |
| MAAP-C02-S03-internal | 457 | 0.046 | 21 |
| MAAP-C02-S03-lapel | 457 | 0.028 | 13 |
| MAAP-C03-S01-internal | 904 | 0.024 | 22 |
| MAAP-C03-S01-lapel | 904 | 0.019 | 17 |
| MAAP-C03-S02-internal | 552 | 0.045 | 25 |
| MAAP-C03-S02-lapel | 552 | 0.016 | 9 |
| MAAP-C03-S03-internal | 600 | 0.028 | 17 |
| MAAP-C03-S03-lapel | 600 | 0.042 | 25 |
| MAAP-C03-S04-internal | 596 | 0.037 | 22 |
| MAAP-C03-S04-lapel | 596 | 0.029 | 17 |
| MAAP-C03-S05-internal | 686 | 0.026 | 18 |
| MAAP-C03-S05-lapel | 686 | 0.032 | 22 |
| MAAP-C03-S06-internal | 507 | 0.039 | 20 |
| MAAP-C03-S06-lapel | 507 | 0.036 | 18 |
| MAAP-C03-S07-internal | 638 | 0.022 | 14 |
| MAAP-C03-S07-lapel | 638 | 0.011 | 7 |
| MAAP-C03-S08-internal | 619 | 0.061 | 38 |
| MAAP-C03-S08-lapel | 619 | 0.074 | 46 |
| MAAP-C03-S10-internal | 789 | 0.023 | 18 |
| MAAP-C03-S10-lapel | 789 | 0.018 | 14 |
| MAAP-C04-S01-internal | 854 | 0.012 | 10 |
| MAAP-C04-S01-lapel | 854 | 0.027 | 23 |
| MAAP-C04-S02-internal | 453 | 0.042 | 19 |
| MAAP-C04-S02-lapel | 453 | 0.033 | 15 |
| MAAP-C04-S03-internal | 578 | 0.043 | 25 |
| MAAP-C04-S03-lapel | 578 | 0.022 | 13 |
| MAAP-C04-S04-internal | 665 | 0.024 | 16 |
| MAAP-C04-S04-lapel | 665 | 0.038 | 25 |
| MAAP-C04-S06-internal | 697 | 0.047 | 33 |
| MAAP-C04-S06-lapel | 697 | 0.044 | 31 |
| MAAP-C04-S07-internal | 693 | 0.029 | 20 |
| MAAP-C04-S07-lapel | 693 | 0.020 | 14 |
| MAAP-C04-S08-internal | 548 | 0.016 | 9 |
| MAAP-C04-S08-lapel | 548 | 0.026 | 14 |
| MAAP-C04-S10-internal | 643 | 0.054 | 35 |
| MAAP-C04-S10-lapel | 643 | 0.033 | 21 |
| MAAP-C05-S01-internal | 655 | 0.034 | 22 |
| MAAP-C05-S01-lapel | 655 | 0.029 | 19 |
| MAAP-C05-S02-internal | 742 | 0.039 | 29 |
| MAAP-C05-S02-lapel | 742 | 0.020 | 15 |
| MAAP-C05-S03-internal | 610 | 0.021 | 13 |
| MAAP-C05-S03-lapel | 610 | 0.016 | 10 |
| MAAP-C05-S04-internal | 773 | 0.023 | 18 |
| MAAP-C05-S04-lapel | 773 | 0.018 | 14 |
| MAAP-C05-S05-internal | 560 | 0.066 | 37 |
| MAAP-C05-S05-lapel | 560 | 0.055 | 31 |
| MAAP-C06-S01-internal | 681 | 0.059 | 40 |
| MAAP-C06-S01-lapel | 681 | 0.053 | 36 |
| MAAP-C06-S02-internal | 719 | 0.031 | 22 |
| MAAP-C06-S02-lapel | 719 | 0.038 | 27 |
| MAAP-C06-S03-internal | 683 | 0.044 | 30 |
| MAAP-C06-S03-lapel | 683 | 0.012 | 8 |
| MAAP-C06-S04-internal | 751 | 0.027 | 20 |
| MAAP-C06-S04-lapel | 751 | 0.025 | 19 |
| MAAP-C07-S01-internal | 161 | 0.075 | 12 |
| MAAP-C07-S01-lapel | 161 | 0.050 | 8 |
| MAAP-C07-S02-internal | 700 | 0.063 | 44 |
| MAAP-C07-S02-lapel | 700 | 0.027 | 19 |
| MAAP-C07-S03-internal | 689 | 0.033 | 23 |
| MAAP-C07-S03-lapel | 689 | 0.016 | 11 |
| MAAP-C07-S04-lapel | 620 | 0.042 | 26 |
| MAAP-C07-S05-internal | 806 | 0.066 | 53 |
| MAAP-C07-S05-lapel | 806 | 0.026 | 21 |
| MAAP-C07-S06-internal | 766 | 0.042 | 32 |
| MAAP-C07-S06-lapel | 766 | 0.030 | 23 |
| MAAP-C07-S07-internal | 765 | 0.031 | 24 |
| MAAP-C07-S07-lapel | 765 | 0.018 | 14 |
| MAAP-C08-S01-internal | 437 | 0.034 | 15 |
| MAAP-C08-S01-lapel | 437 | 0.018 | 8 |
| MAAP-C08-S02-internal | 743 | 0.038 | 28 |
| MAAP-C08-S02-lapel | 743 | 0.023 | 22 |
| MAAP-C08-S03-internal | 463 | 0.028 | 13 |
| MAAP-C08-S03-lapel | 463 | 0.041 | 19 |
| MAAP-C08-S04-internal | 1633 | 0.010 | 16 |
| MAAP-C08-S04-lapel | 1633 | 0.017 | 28 |
| MAAP-C08-S05-internal | 456 | 0.029 | 13 |
| MAAP-C08-S05-lapel | 456 | 0.035 | 16 |
| MAAP-C08-S06-internal | 634 | 0.039 | 25 |
| MAAP-C08-S06-lapel | 634 | 0.038 | 24 |
| MAAP-C08-S07-internal | 680 | 0.037 | 25 |
| MAAP-C08-S07-lapel | 680 | 0.040 | 27 |
| MAAP-C08-S08-internal | 510 | 0.014 | 7 |
| MAAP-C08-S08-lapel | 510 | 0.020 | 10 |
| MAAP-C08-S09-lapel | 684 | 0.038 | 26 |
| MAAP-C08-S10-internal | 578 | 0.036 | 21 |
| MAAP-C08-S10-lapel | 578 | 0.022 | 13 |
| MAAP-C08-S11-internal | 532 | 0.024 | 13 |
| MAAP-C08-S11-lapel | 532 | 0.028 | 15 |
| MAAP-C09-S01-internal | 634 | 0.041 | 26 |
| MAAP-C09-S01-lapel | 634 | 0.009 | 6 |
| MAAP-C09-S02-internal | 667 | 0.027 | 18 |
| MAAP-C09-S02-lapel | 667 | 0.067 | 45 |
| MAAP-C09-S03-internal | 584 | 0.027 | 16 |
| MAAP-C09-S03-lapel | 584 | 0.026 | 15 |
| MAAP-C09-S04-internal | 572 | 0.021 | 12 |
| MAAP-C09-S04-lapel | 572 | 0.035 | 20 |
| MAAP-C09-S05-internal | 552 | 0.042 | 23 |
| MAAP-C09-S05-lapel | 552 | 0.011 | 6 |
| MAAP-C09-S06-internal | 636 | 0.031 | 20 |
| MAAP-C09-S06-lapel | 636 | 0.044 | 28 |
| MAAP-C09-S07-lapel | 688 | 0.025 | 17 |
| MAAP-C09-S08-internal | 616 | 0.068 | 42 |
| MAAP-C09-S08-lapel | 616 | 0.013 | 8 |
| MAAP-C09-S09-internal | 589 | 0.048 | 28 |
| MAAP-C09-S09-lapel | 589 | 0.014 | 8 |
| MAAP-C09-S10-internal | 687 | 0.055 | 38 |
| MAAP-C09-S10-lapel | 687 | 0.047 | 32 |
| MAAP-C10-S01-internal | 541 | 0.050 | 27 |
| MAAP-C10-S01-lapel | 541 | 0.018 | 10 |
| MAAP-C10-S02-internal | 385 | 0.057 | 22 |
| MAAP-C10-S02-lapel | 385 | 0.034 | 13 |
| MAAP-C10-S03-internal | 519 | 0.025 | 13 |
| MAAP-C10-S03-lapel | 519 | 0.037 | 19 |
| MAAP-C10-S04-internal | 699 | 0.034 | 24 |
| MAAP-C10-S04-lapel | 699 | 0.043 | 30 |
| MAAP-C10-S05-internal | 665 | 0.047 | 31 |
| MAAP-C10-S05-lapel | 665 | 0.033 | 22 |
| MAAP-C10-S06-internal | 738 | 0.069 | 51 |
| MAAP-C10-S06-lapel | 738 | 0.045 | 33 |
| MAAP-C10-S07-lapel | 638 | 0.044 | 28 |
| MAAP-C10-S08-internal | 801 | 0.039 | 31 |
| MAAP-C10-S08-lapel | 801 | 0.022 | 18 |
| MAAP-C10-S09-internal | 495 | 0.030 | 15 |
| MAAP-C10-S09-lapel | 495 | 0.034 | 17 |
| MAAP-C11-S01-internal | 220 | 0.027 | 6 |
| MAAP-C11-S01-lapel | 220 | 0.009 | 2 |
| MAAP-C11-S02-internal | 299 | 0.043 | 13 |
| MAAP-C11-S02-lapel | 299 | 0.033 | 10 |
| MAAP-C3-S09-internal | 765 | 0.047 | 36 |
| MAAP-C3-S09-lapel | 765 | 0.014 | 11 |
| MAAP-C4-S9-internal | 726 | 0.062 | 45 |
| MAAP-C4-S9-lapel | 726 | 0.026 | 19 |
| UH-C03-S01-internal | 596 | 0.017 | 10 |
| UH-C03-S01-lapel | 596 | 0.012 | 7 |
| UH-C04-S01-internal | 266 | 0.049 | 13 |
| UH-C04-S01-lapel | 266 | 0.019 | 5 |
| UH-C05-S01-internal | 555 | 0.022 | 12 |
| UH-C05-S01-lapel | 555 | 0.022 | 12 |
| UH-C06-S01-internal | 833 | 0.047 | 39 |
| UH-C06-S01-lapel | 833 | 0.035 | 29 |
| UH-C01-S02-internal | 883 | 0.080 | 71 |
| UH-C01-S02-lapel | 883 | 0.104 | 92 |
| UH-C04-S02-internal | 743 | 0.032 | 24 |
| UH-C04-S02-lapel | 743 | 0.082 | 61 |
| UH-C05-S02-internal | 545 | 0.057 | 31 |
| UH-C05-S02-lapel | 545 | 0.064 | 35 |
| UH-C01-S03-internal | 731 | 0.100 | 73 |
| UH-C01-S03-lapel | 731 | 0.073 | 53 |
| UH-C03-S02-internal | 525 | 0.021 | 11 |
| UH-C03-S02-lapel | 525 | 0.023 | 12 |
| UH-C04-S03-internal | 600 | 0.033 | 20 |
| UH-C04-S03-lapel | 600 | 0.025 | 15 |
| UH-C05-S03-internal | 93 | 0.022 | 2 |
| UH-C05-S03-lapel | 93 | 0.022 | 2 |
| UH-C01-S04-internal | 718 | 0.058 | 42 |
| UH-C01-S04-lapel | 718 | 0.043 | 31 |
| UH-C03-S03-internal | 640 | 0.020 | 13 |
| UH-C03-S03-lapel | 640 | 0.014 | 9 |
| UH-C04-S04-internal | 357 | 0.025 | 9 |
| UH-C04-S04-lapel | 356 | 0.011 | 4 |
| UH-C05-S04-internal | 634 | 0.032 | 20 |
| UH-C05-S04-lapel | 634 | 0.016 | 10 |
| UH-C07-S01-internal | 612 | 0.028 | 17 |
| UH-C07-S01-lapel | 612 | 0.014 | 9 |

**Table A.2:**
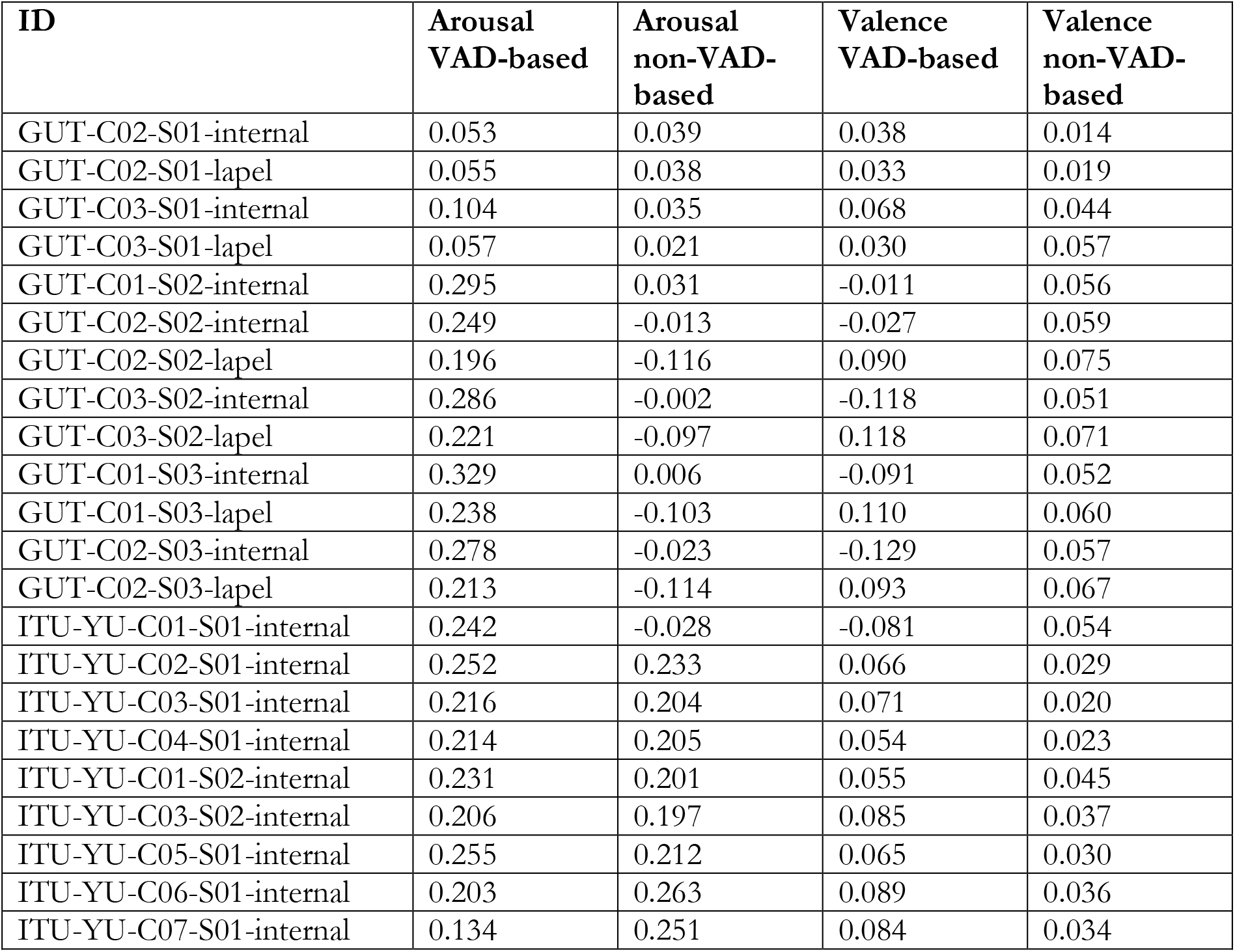
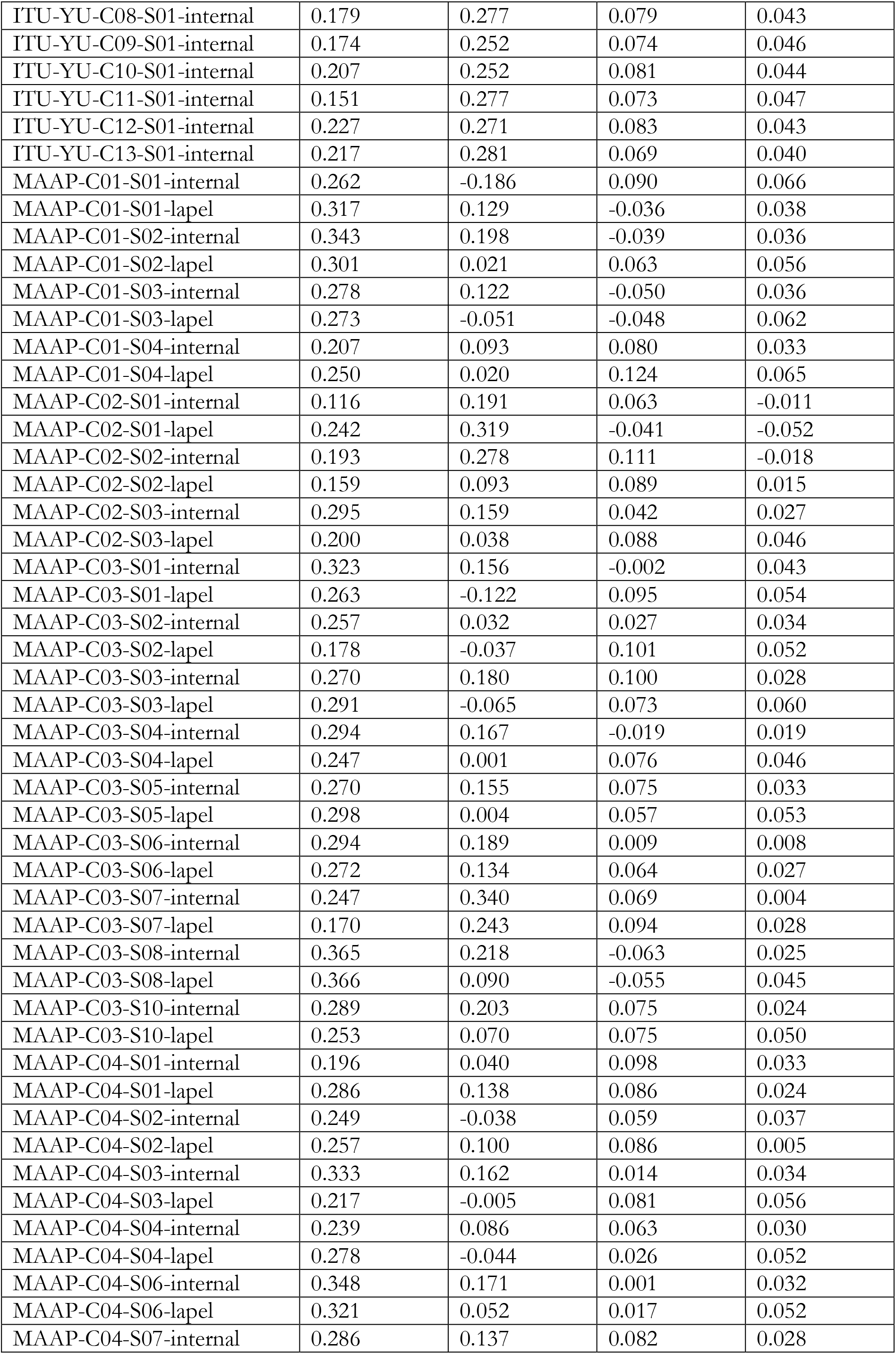
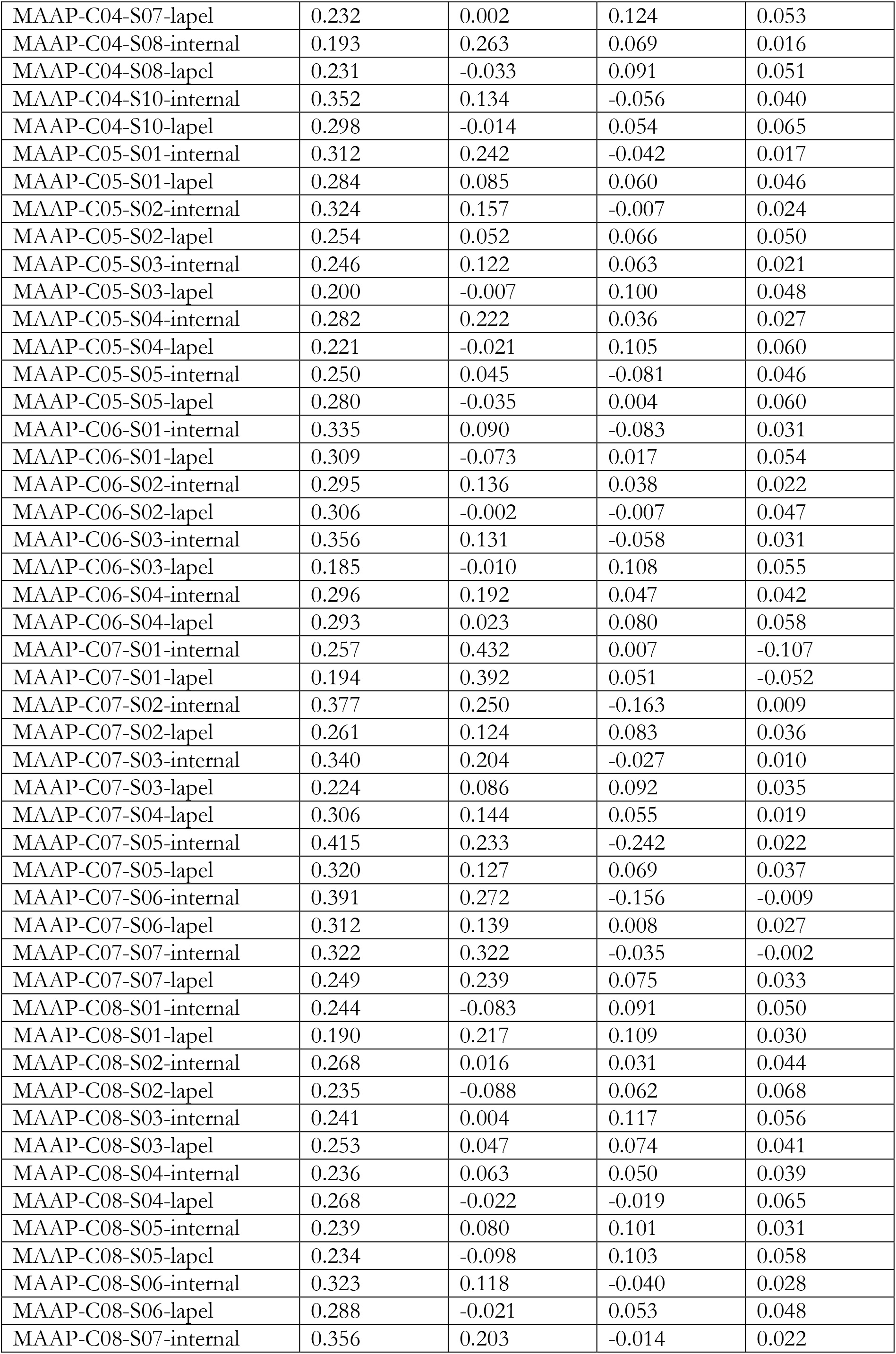
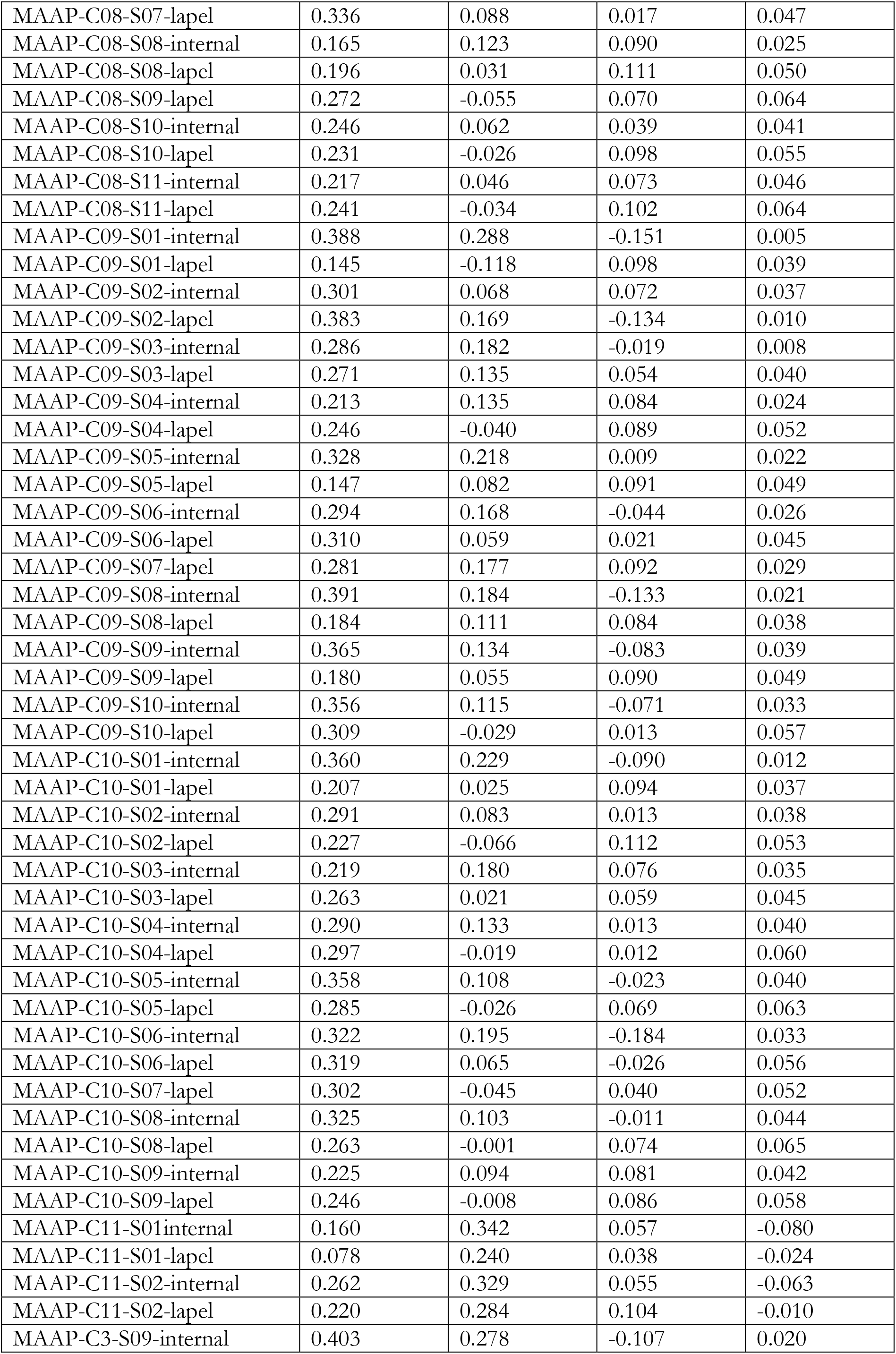
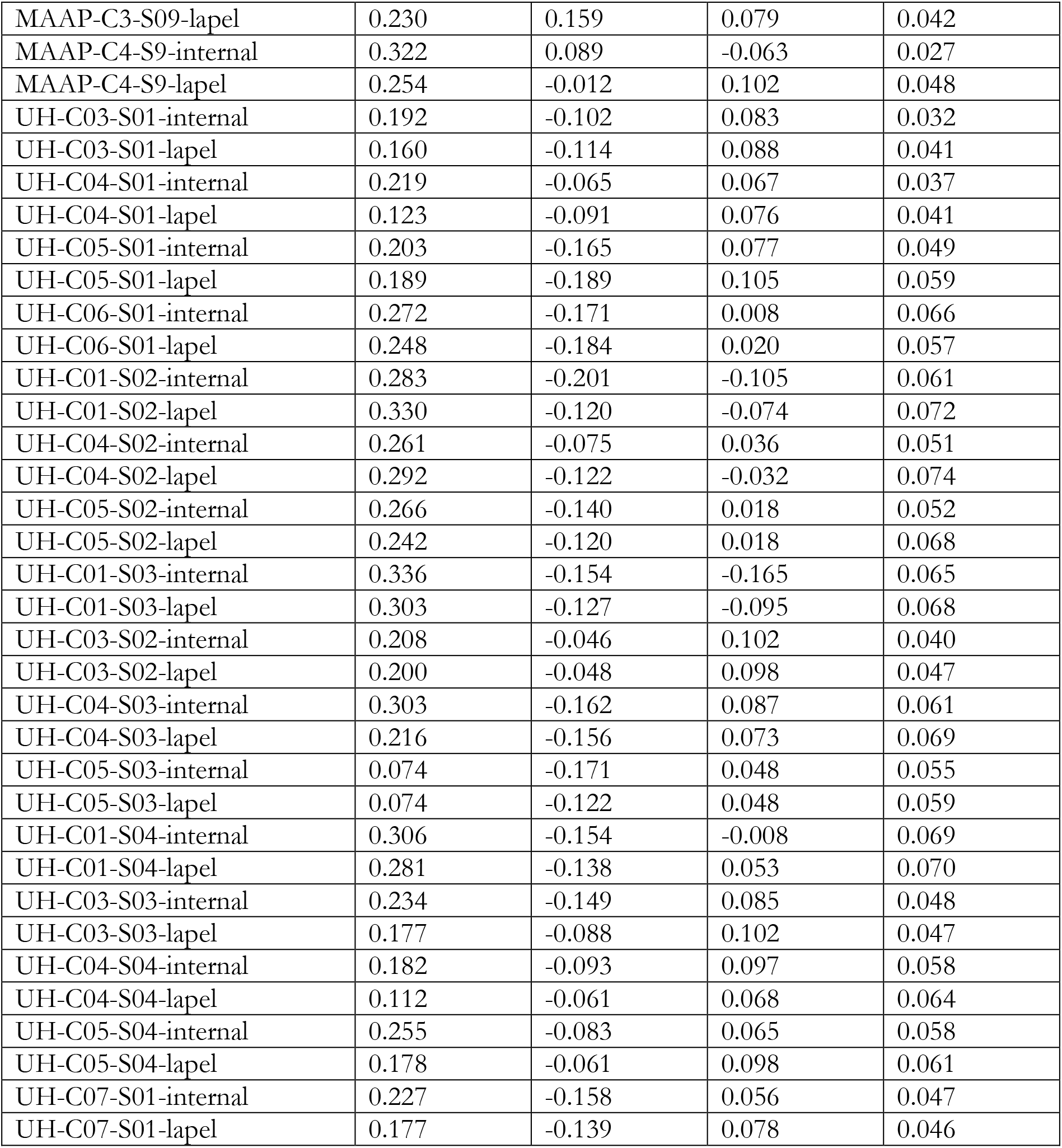
Average arousal and valence values per intervention session. GUT = Gdansk University of Technology, Poland; ITU-YU = Istanbul Teknik Universitesi and Yeditepe University Vakif, Turkey; MAAP = Macedonian Association for Applied Psychology, North Macedonia; non-VAD-based = based on the whole audio material; UH = University of Hertfordshire, United Kingdom; VAD-based = based on all detected child vocalisations.

| **ID** | **Arousal**<br>**VAD-based** | **Arousal**<br>**non-VAD-**<br>**based** | **Valence**<br>**VAD-based** | **Valence**<br>**non-VAD-**<br>**based** |
| --- | --- | --- | --- | --- |
| GUT-C02-S01-internal | 0.053 | 0.039 | 0.038 | 0.014 |
| GUT-C02-S01-lapel | 0.055 | 0.038 | 0.033 | 0.019 |
| GUT-C03-S01-internal | 0.104 | 0.035 | 0.068 | 0.044 |
| GUT-C03-S01-lapel | 0.057 | 0.021 | 0.030 | 0.057 |
| GUT-C01-S02-internal | 0.295 | 0.031 | -0.011 | 0.056 |
| GUT-C02-S02-internal | 0.249 | -0.013 | -0.027 | 0.059 |
| GUT-C02-S02-lapel | 0.196 | -0.116 | 0.090 | 0.075 |
| GUT-C03-S02-internal | 0.286 | -0.002 | -0.118 | 0.051 |
| GUT-C03-S02-lapel | 0.221 | -0.097 | 0.118 | 0.071 |
| GUT-C01-S03-internal | 0.329 | 0.006 | -0.091 | 0.052 |
| GUT-C01-S03-lapel | 0.238 | -0.103 | 0.110 | 0.060 |
| GUT-C02-S03-internal | 0.278 | -0.023 | -0.129 | 0.057 |
| GUT-C02-S03-lapel | 0.213 | -0.114 | 0.093 | 0.067 |
| ITU-YU-C01-S01-internal | 0.242 | -0.028 | -0.081 | 0.054 |
| ITU-YU-C02-S01-internal | 0.252 | 0.233 | 0.066 | 0.029 |
| ITU-YU-C03-S01-internal | 0.216 | 0.204 | 0.071 | 0.020 |
| ITU-YU-C04-S01-internal | 0.214 | 0.205 | 0.054 | 0.023 |
| ITU-YU-C01-S02-internal | 0.231 | 0.201 | 0.055 | 0.045 |
| ITU-YU-C03-S02-internal | 0.206 | 0.197 | 0.085 | 0.037 |
| ITU-YU-C05-S01-internal | 0.255 | 0.212 | 0.065 | 0.030 |
| ITU-YU-C06-S01-internal | 0.203 | 0.263 | 0.089 | 0.036 |
| ITU-YU-C07-S01-internal | 0.134 | 0.251 | 0.084 | 0.034 |
| ITU-YU-C08-S01-internal | 0.179 | 0.277 | 0.079 | 0.043 |
| ITU-YU-C09-S01-internal | 0.174 | 0.252 | 0.074 | 0.046 |
| ITU-YU-C10-S01-internal | 0.207 | 0.252 | 0.081 | 0.044 |
| ITU-YU-C11-S01-internal | 0.151 | 0.277 | 0.073 | 0.047 |
| ITU-YU-C12-S01-internal | 0.227 | 0.271 | 0.083 | 0.043 |
| ITU-YU-C13-S01-internal | 0.217 | 0.281 | 0.069 | 0.040 |
| MAAP-C01-S01-internal | 0.262 | -0.186 | 0.090 | 0.066 |
| MAAP-C01-S01-lapel | 0.317 | 0.129 | -0.036 | 0.038 |
| MAAP-C01-S02-internal | 0.343 | 0.198 | -0.039 | 0.036 |
| MAAP-C01-S02-lapel | 0.301 | 0.021 | 0.063 | 0.056 |
| MAAP-C01-S03-internal | 0.278 | 0.122 | -0.050 | 0.036 |
| MAAP-C01-S03-lapel | 0.273 | -0.051 | -0.048 | 0.062 |
| MAAP-C01-S04-internal | 0.207 | 0.093 | 0.080 | 0.033 |
| MAAP-C01-S04-lapel | 0.250 | 0.020 | 0.124 | 0.065 |
| MAAP-C02-S01-internal | 0.116 | 0.191 | 0.063 | -0.011 |
| MAAP-C02-S01-lapel | 0.242 | 0.319 | -0.041 | -0.052 |
| MAAP-C02-S02-internal | 0.193 | 0.278 | 0.111 | -0.018 |
| MAAP-C02-S02-lapel | 0.159 | 0.093 | 0.089 | 0.015 |
| MAAP-C02-S03-internal | 0.295 | 0.159 | 0.042 | 0.027 |
| MAAP-C02-S03-lapel | 0.200 | 0.038 | 0.088 | 0.046 |
| MAAP-C03-S01-internal | 0.323 | 0.156 | -0.002 | 0.043 |
| MAAP-C03-S01-lapel | 0.263 | -0.122 | 0.095 | 0.054 |
| MAAP-C03-S02-internal | 0.257 | 0.032 | 0.027 | 0.034 |
| MAAP-C03-S02-lapel | 0.178 | -0.037 | 0.101 | 0.052 |
| MAAP-C03-S03-internal | 0.270 | 0.180 | 0.100 | 0.028 |
| MAAP-C03-S03-lapel | 0.291 | -0.065 | 0.073 | 0.060 |
| MAAP-C03-S04-internal | 0.294 | 0.167 | -0.019 | 0.019 |
| MAAP-C03-S04-lapel | 0.247 | 0.001 | 0.076 | 0.046 |
| MAAP-C03-S05-internal | 0.270 | 0.155 | 0.075 | 0.033 |
| MAAP-C03-S05-lapel | 0.298 | 0.004 | 0.057 | 0.053 |
| MAAP-C03-S06-internal | 0.294 | 0.189 | 0.009 | 0.008 |
| MAAP-C03-S06-lapel | 0.272 | 0.134 | 0.064 | 0.027 |
| MAAP-C03-S07-internal | 0.247 | 0.340 | 0.069 | 0.004 |
| MAAP-C03-S07-lapel | 0.170 | 0.243 | 0.094 | 0.028 |
| MAAP-C03-S08-internal | 0.365 | 0.218 | -0.063 | 0.025 |
| MAAP-C03-S08-lapel | 0.366 | 0.090 | -0.055 | 0.045 |
| MAAP-C03-S10-internal | 0.289 | 0.203 | 0.075 | 0.024 |
| MAAP-C03-S10-lapel | 0.253 | 0.070 | 0.075 | 0.050 |
| MAAP-C04-S01-internal | 0.196 | 0.040 | 0.098 | 0.033 |
| MAAP-C04-S01-lapel | 0.286 | 0.138 | 0.086 | 0.024 |
| MAAP-C04-S02-internal | 0.249 | -0.038 | 0.059 | 0.037 |
| MAAP-C04-S02-lapel | 0.257 | 0.100 | 0.086 | 0.005 |
| MAAP-C04-S03-internal | 0.333 | 0.162 | 0.014 | 0.034 |
| MAAP-C04-S03-lapel | 0.217 | -0.005 | 0.081 | 0.056 |
| MAAP-C04-S04-internal | 0.239 | 0.086 | 0.063 | 0.030 |
| MAAP-C04-S04-lapel | 0.278 | -0.044 | 0.026 | 0.052 |
| MAAP-C04-S06-internal | 0.348 | 0.171 | 0.001 | 0.032 |
| MAAP-C04-S06-lapel | 0.321 | 0.052 | 0.017 | 0.052 |
| MAAP-C04-S07-internal | 0.286 | 0.137 | 0.082 | 0.028 |
| MAAP-C04-S07-lapel | 0.232 | 0.002 | 0.124 | 0.053 |
| MAAP-C04-S08-internal | 0.193 | 0.263 | 0.069 | 0.016 |
| MAAP-C04-S08-lapel | 0.231 | -0.033 | 0.091 | 0.051 |
| MAAP-C04-S10-internal | 0.352 | 0.134 | -0.056 | 0.040 |
| MAAP-C04-S10-lapel | 0.298 | -0.014 | 0.054 | 0.065 |
| MAAP-C05-S01-internal | 0.312 | 0.242 | -0.042 | 0.017 |
| MAAP-C05-S01-lapel | 0.284 | 0.085 | 0.060 | 0.046 |
| MAAP-C05-S02-internal | 0.324 | 0.157 | -0.007 | 0.024 |
| MAAP-C05-S02-lapel | 0.254 | 0.052 | 0.066 | 0.050 |
| MAAP-C05-S03-internal | 0.246 | 0.122 | 0.063 | 0.021 |
| MAAP-C05-S03-lapel | 0.200 | -0.007 | 0.100 | 0.048 |
| MAAP-C05-S04-internal | 0.282 | 0.222 | 0.036 | 0.027 |
| MAAP-C05-S04-lapel | 0.221 | -0.021 | 0.105 | 0.060 |
| MAAP-C05-S05-internal | 0.250 | 0.045 | -0.081 | 0.046 |
| MAAP-C05-S05-lapel | 0.280 | -0.035 | 0.004 | 0.060 |
| MAAP-C06-S01-internal | 0.335 | 0.090 | -0.083 | 0.031 |
| MAAP-C06-S01-lapel | 0.309 | -0.073 | 0.017 | 0.054 |
| MAAP-C06-S02-internal | 0.295 | 0.136 | 0.038 | 0.022 |
| MAAP-C06-S02-lapel | 0.306 | -0.002 | -0.007 | 0.047 |
| MAAP-C06-S03-internal | 0.356 | 0.131 | -0.058 | 0.031 |
| MAAP-C06-S03-lapel | 0.185 | -0.010 | 0.108 | 0.055 |
| MAAP-C06-S04-internal | 0.296 | 0.192 | 0.047 | 0.042 |
| MAAP-C06-S04-lapel | 0.293 | 0.023 | 0.080 | 0.058 |
| MAAP-C07-S01-internal | 0.257 | 0.432 | 0.007 | -0.107 |
| MAAP-C07-S01-lapel | 0.194 | 0.392 | 0.051 | -0.052 |
| MAAP-C07-S02-internal | 0.377 | 0.250 | -0.163 | 0.009 |
| MAAP-C07-S02-lapel | 0.261 | 0.124 | 0.083 | 0.036 |
| MAAP-C07-S03-internal | 0.340 | 0.204 | -0.027 | 0.010 |
| MAAP-C07-S03-lapel | 0.224 | 0.086 | 0.092 | 0.035 |
| MAAP-C07-S04-lapel | 0.306 | 0.144 | 0.055 | 0.019 |
| MAAP-C07-S05-internal | 0.415 | 0.233 | -0.242 | 0.022 |
| MAAP-C07-S05-lapel | 0.320 | 0.127 | 0.069 | 0.037 |
| MAAP-C07-S06-internal | 0.391 | 0.272 | -0.156 | -0.009 |
| MAAP-C07-S06-lapel | 0.312 | 0.139 | 0.008 | 0.027 |
| MAAP-C07-S07-internal | 0.322 | 0.322 | -0.035 | -0.002 |
| MAAP-C07-S07-lapel | 0.249 | 0.239 | 0.075 | 0.033 |
| MAAP-C08-S01-internal | 0.244 | -0.083 | 0.091 | 0.050 |
| MAAP-C08-S01-lapel | 0.190 | 0.217 | 0.109 | 0.030 |
| MAAP-C08-S02-internal | 0.268 | 0.016 | 0.031 | 0.044 |
| MAAP-C08-S02-lapel | 0.235 | -0.088 | 0.062 | 0.068 |
| MAAP-C08-S03-internal | 0.241 | 0.004 | 0.117 | 0.056 |
| MAAP-C08-S03-lapel | 0.253 | 0.047 | 0.074 | 0.041 |
| MAAP-C08-S04-internal | 0.236 | 0.063 | 0.050 | 0.039 |
| MAAP-C08-S04-lapel | 0.268 | -0.022 | -0.019 | 0.065 |
| MAAP-C08-S05-internal | 0.239 | 0.080 | 0.101 | 0.031 |
| MAAP-C08-S05-lapel | 0.234 | -0.098 | 0.103 | 0.058 |
| MAAP-C08-S06-internal | 0.323 | 0.118 | -0.040 | 0.028 |
| MAAP-C08-S06-lapel | 0.288 | -0.021 | 0.053 | 0.048 |
| MAAP-C08-S07-internal | 0.356 | 0.203 | -0.014 | 0.022 |
| MAAP-C08-S07-lapel | 0.336 | 0.088 | 0.017 | 0.047 |
| MAAP-C08-S08-internal | 0.165 | 0.123 | 0.090 | 0.025 |
| MAAP-C08-S08-lapel | 0.196 | 0.031 | 0.111 | 0.050 |
| MAAP-C08-S09-lapel | 0.272 | -0.055 | 0.070 | 0.064 |
| MAAP-C08-S10-internal | 0.246 | 0.062 | 0.039 | 0.041 |
| MAAP-C08-S10-lapel | 0.231 | -0.026 | 0.098 | 0.055 |
| MAAP-C08-S11-internal | 0.217 | 0.046 | 0.073 | 0.046 |
| MAAP-C08-S11-lapel | 0.241 | -0.034 | 0.102 | 0.064 |
| MAAP-C09-S01-internal | 0.388 | 0.288 | -0.151 | 0.005 |
| MAAP-C09-S01-lapel | 0.145 | -0.118 | 0.098 | 0.039 |
| MAAP-C09-S02-internal | 0.301 | 0.068 | 0.072 | 0.037 |
| MAAP-C09-S02-lapel | 0.383 | 0.169 | -0.134 | 0.010 |
| MAAP-C09-S03-internal | 0.286 | 0.182 | -0.019 | 0.008 |
| MAAP-C09-S03-lapel | 0.271 | 0.135 | 0.054 | 0.040 |
| MAAP-C09-S04-internal | 0.213 | 0.135 | 0.084 | 0.024 |
| MAAP-C09-S04-lapel | 0.246 | -0.040 | 0.089 | 0.052 |
| MAAP-C09-S05-internal | 0.328 | 0.218 | 0.009 | 0.022 |
| MAAP-C09-S05-lapel | 0.147 | 0.082 | 0.091 | 0.049 |
| MAAP-C09-S06-internal | 0.294 | 0.168 | -0.044 | 0.026 |
| MAAP-C09-S06-lapel | 0.310 | 0.059 | 0.021 | 0.045 |
| MAAP-C09-S07-lapel | 0.281 | 0.177 | 0.092 | 0.029 |
| MAAP-C09-S08-internal | 0.391 | 0.184 | -0.133 | 0.021 |
| MAAP-C09-S08-lapel | 0.184 | 0.111 | 0.084 | 0.038 |
| MAAP-C09-S09-internal | 0.365 | 0.134 | -0.083 | 0.039 |
| MAAP-C09-S09-lapel | 0.180 | 0.055 | 0.090 | 0.049 |
| MAAP-C09-S10-internal | 0.356 | 0.115 | -0.071 | 0.033 |
| MAAP-C09-S10-lapel | 0.309 | -0.029 | 0.013 | 0.057 |
| MAAP-C10-S01-internal | 0.360 | 0.229 | -0.090 | 0.012 |
| MAAP-C10-S01-lapel | 0.207 | 0.025 | 0.094 | 0.037 |
| MAAP-C10-S02-internal | 0.291 | 0.083 | 0.013 | 0.038 |
| MAAP-C10-S02-lapel | 0.227 | -0.066 | 0.112 | 0.053 |
| MAAP-C10-S03-internal | 0.219 | 0.180 | 0.076 | 0.035 |
| MAAP-C10-S03-lapel | 0.263 | 0.021 | 0.059 | 0.045 |
| MAAP-C10-S04-internal | 0.290 | 0.133 | 0.013 | 0.040 |
| MAAP-C10-S04-lapel | 0.297 | -0.019 | 0.012 | 0.060 |
| MAAP-C10-S05-internal | 0.358 | 0.108 | -0.023 | 0.040 |
| MAAP-C10-S05-lapel | 0.285 | -0.026 | 0.069 | 0.063 |
| MAAP-C10-S06-internal | 0.322 | 0.195 | -0.184 | 0.033 |
| MAAP-C10-S06-lapel | 0.319 | 0.065 | -0.026 | 0.056 |
| MAAP-C10-S07-lapel | 0.302 | -0.045 | 0.040 | 0.052 |
| MAAP-C10-S08-internal | 0.325 | 0.103 | -0.011 | 0.044 |
| MAAP-C10-S08-lapel | 0.263 | -0.001 | 0.074 | 0.065 |
| MAAP-C10-S09-internal | 0.225 | 0.094 | 0.081 | 0.042 |
| MAAP-C10-S09-lapel | 0.246 | -0.008 | 0.086 | 0.058 |
| MAAP-C11-S01-internal | 0.160 | 0.342 | 0.057 | -0.080 |
| MAAP-C11-S01-lapel | 0.078 | 0.240 | 0.038 | -0.024 |
| MAAP-C11-S02-internal | 0.262 | 0.329 | 0.055 | -0.063 |
| MAAP-C11-S02-lapel | 0.220 | 0.284 | 0.104 | -0.010 |
| MAAP-C3-S09-internal | 0.403 | 0.278 | -0.107 | 0.020 |
| MAAP-C3-S09-lapel | 0.230 | 0.159 | 0.079 | 0.042 |
| MAAP-C4-S9-internal | 0.322 | 0.089 | -0.063 | 0.027 |
| MAAP-C4-S9-lapel | 0.254 | -0.012 | 0.102 | 0.048 |
| UH-C03-S01-internal | 0.192 | -0.102 | 0.083 | 0.032 |
| UH-C03-S01-lapel | 0.160 | -0.114 | 0.088 | 0.041 |
| UH-C04-S01-internal | 0.219 | -0.065 | 0.067 | 0.037 |
| UH-C04-S01-lapel | 0.123 | -0.091 | 0.076 | 0.041 |
| UH-C05-S01-internal | 0.203 | -0.165 | 0.077 | 0.049 |
| UH-C05-S01-lapel | 0.189 | -0.189 | 0.105 | 0.059 |
| UH-C06-S01-internal | 0.272 | -0.171 | 0.008 | 0.066 |
| UH-C06-S01-lapel | 0.248 | -0.184 | 0.020 | 0.057 |
| UH-C01-S02-internal | 0.283 | -0.201 | -0.105 | 0.061 |
| UH-C01-S02-lapel | 0.330 | -0.120 | -0.074 | 0.072 |
| UH-C04-S02-internal | 0.261 | -0.075 | 0.036 | 0.051 |
| UH-C04-S02-lapel | 0.292 | -0.122 | -0.032 | 0.074 |
| UH-C05-S02-internal | 0.266 | -0.140 | 0.018 | 0.052 |
| UH-C05-S02-lapel | 0.242 | -0.120 | 0.018 | 0.068 |
| UH-C01-S03-internal | 0.336 | -0.154 | -0.165 | 0.065 |
| UH-C01-S03-lapel | 0.303 | -0.127 | -0.095 | 0.068 |
| UH-C03-S02-internal | 0.208 | -0.046 | 0.102 | 0.040 |
| UH-C03-S02-lapel | 0.200 | -0.048 | 0.098 | 0.047 |
| UH-C04-S03-internal | 0.303 | -0.162 | 0.087 | 0.061 |
| UH-C04-S03-lapel | 0.216 | -0.156 | 0.073 | 0.069 |
| UH-C05-S03-internal | 0.074 | -0.171 | 0.048 | 0.055 |
| UH-C05-S03-lapel | 0.074 | -0.122 | 0.048 | 0.059 |
| UH-C01-S04-internal | 0.306 | -0.154 | -0.008 | 0.069 |
| UH-C01-S04-lapel | 0.281 | -0.138 | 0.053 | 0.070 |
| UH-C03-S03-internal | 0.234 | -0.149 | 0.085 | 0.048 |
| UH-C03-S03-lapel | 0.177 | -0.088 | 0.102 | 0.047 |
| UH-C04-S04-internal | 0.182 | -0.093 | 0.097 | 0.058 |
| UH-C04-S04-lapel | 0.112 | -0.061 | 0.068 | 0.064 |
| UH-C05-S04-internal | 0.255 | -0.083 | 0.065 | 0.058 |
| UH-C05-S04-lapel | 0.178 | -0.061 | 0.098 | 0.061 |
| UH-C07-S01-internal | 0.227 | -0.158 | 0.056 | 0.047 |
| UH-C07-S01-lapel | 0.177 | -0.139 | 0.078 | 0.046 |

